# Interpretable Phenotype Discovery Using Temporal Risk-Aware Clustering

**DOI:** 10.64898/2026.10.18.26365169

**Authors:** Asal Roudbari, Pascal Van Hentenryck, Reza Zandehshahvar

## Abstract

Longitudinal clinical cohorts contain heterogeneous progression patterns that are obscured by coarse diagnostic labels. Discovering these latent progression phenotypes requires models that distinguish patients not only by their outcomes, but also by how their clinical trajectories evolve. This paper proposes TRACE, an interpretable temporal phenotyping method that enables the learning of outcome-informed progression phenotypes from longitudinal trajectories. TRACE combines a trajectory-and outcome-aware contrastive objective with a probabilistic predictive phenotype layer, encouraging phenotypes to capture progression structure while remaining predictive of diagnostic state. A dual-attention mechanism further supports interpretation by identifying the visits and clinical variables most associated with each phenotype assignment. TRACE is evaluated on the Alzheimer’s Disease Neuroimaging Initiative (ADNI) cohort for cognitive disease modeling, showing improved performance in both diagnostic prediction and phenotype discovery compared with other temporal phenotyping baselines. Finally, by allowing the number of phenotypes to exceed the diagnostic labels, TRACE enables discovering additional progression patterns that are not captured by labels defined a priori.

## 1 Introduction

Patients sharing a similar diagnosis often follow distinct progression trajectories, differing in progression rate, intermediate physiological states, and treatment response [1]. This heterogeneity carries clinically meaningful information beyond diagnostic labels, motivating methods that discover latent progression mechanisms directly from longitudinal Electronic Health Records (EHRs) [2]. Temporal phenotyping pursues this goal by identifying patient subgroups from longitudinal representations, typically by jointly optimizing phenotype assignments and a predictive objective [3–9]. A central question in temporal phenotyping is therefore *to discover which distinct progression patterns exist within a diagnostic category and whether these patterns are sufficiently predictive to support diagnosis*. [10, 11]

This work introduces TRACE (Temporal Risk-Aware Clustering with Explanations), an interpretable temporal phenotyping framework for longitudinal patient histories. TRACE makes two main contributions: 1) It proposes a new architecture for predictive and interpretable temporal phenotyping, where patient histories are mapped to probabilistic phenotype assignments through a predictive phenotype layer that directly mediates diagnostic prediction. This design is combined with trajectory-and outcome-aware contrastive learning, which encourages the learned phenotypes to reflect both future diagnostic risk and the temporal pathways by which patients reach those outcomes. Dual attention further characterizes each phenotype by identifying the visits and clinical variables most associated with the assignment. 2) TRACE enables phenotype discovery beyond predefined diagnostic labels by allowing the number of latent phenotypes to exceed the number of outcome classes. This flexibility allows multiple progression phenotypes to emerge within the same diagnostic class, revealing heterogeneity that would be obscured by coarse labels defined a priori. TRACE is evaluated on Alzheimer’s disease progression using the Alzheimer’s Disease Neuroimaging Initiative (ADNI) [12], a challenging setting for phenotype discovery because progression is heterogeneous and may unfold gradually through subtle longitudinal changes before overt cognitive decline. Across prediction horizons, TRACE improves over existing temporal phenotyping methods, with the largest gains at the 24-month horizon, where it outperforms the strongest baseline by 3.1 and 5.8 percentage points in Purity and Silhouette Index, respectively.

## 2 Problem formulation

Let a dataset *D* contain *N* patients with longitudinal histories 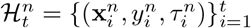, where 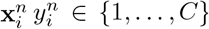, and 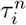 denote the observed features, diagnostic label, and timestamp at visit *i*. The visits may be irregularly spaced in time. For each patient *n*, the objective is to learn a dynamic phenotype representation that captures longitudinal disease heterogeneity while supporting downstream diagnostic prediction. Formally, the goal is to estimate a soft phenotype distribution 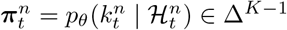, where 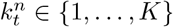 is the latent phenotype assigned to patient *n* at time *t*, and Δ^*K−*1^ the (*K* − 1)-dimensional probability simplex. A phenotype therefore denotes a group of histories that share both a diagnostic outcome and a pattern of longitudinal evolution. Conditioned on the learned patient representation, the secondary goal is to predict the clinical outcome at a future horizon *h* ≥ 0 by estimating 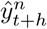. When *h* = 0, the label 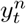 is excluded from the history 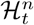. The overall goal is therefore end-to-end phenotype discovery and outcome prediction so that the learned phenotypes remain both clinically interpretable and predictive of disease progression. The superscript *n* is omitted in the following for brevity, unless specified otherwise.

## 3 TRACE

Figure 1 shows the architecture of TRACE, which is composed of four main pillars as follows and detailed in Appendix B.

**Figure 1:**
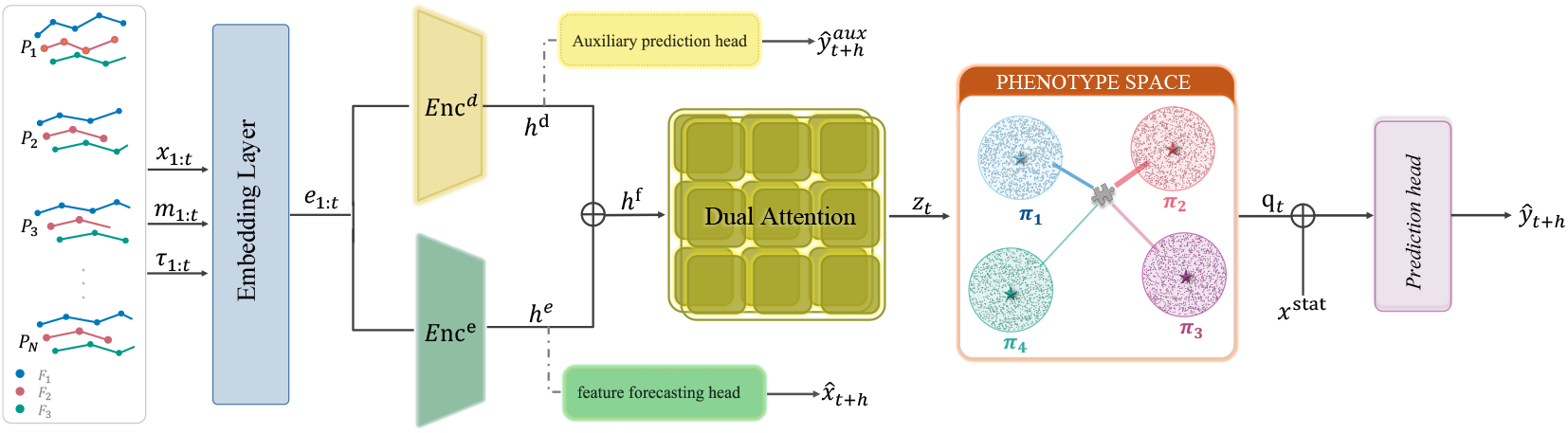
Overview of TRACE. Longitudinal EHR trajectories are embedded and processed by dual transformer encoders that separately model diagnostic state and temporal evolution. Dual attention identifies salient visits and features, and an interpretable phenotype bottleneck maps the resulting context to soft phenotype assignments and the downstream diagnosis.

### Time-aware input embedding (§B.1)

Given the longitudinal data ℋ_*t*_, each visit *i*∈ [1, *t*] is embedded as **e**_*i*_ = Embed(**x**_*i*_, **m**_*i*_, *τ*_*i*_, *δ*_*i*_) using a time-aware embedding layer, where **m**_*i*_ is a missingness indicator mask, and *δ*_*i*_ = *τ*_*i*_ − *τ*_*i−*1_ represents time in between visits. This forms an embedding vector **e**_1:*t*_ ∈ ℝ^*k×t*^.

### Dual-objective encoders (§B.2)

Two encoders provide complementary evidence for phenotype formation: the *diagnostic encoder* captures clinical state information through 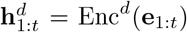, and supervision by the diagnostic label at time *t* + *h*. The *evolution encoder* captures longitudinal trajectories through 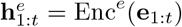 and supervision for predicting the feature values at time *t* + *h*. A mutual-information penalty encourages these streams to capture complementary information by reducing overlap between the diagnostic state and evolution representations. Their fusion 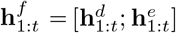 encodes both diagnostic state and the clinical feature trajectories.

### Dual attention and contrastive learning (§B.3)

The encoded vector 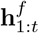 is passed through a dual-attention layer [7], which assigns visit-level weights *α*_*t,i*_ and feature-level weights ***β***_*i*_ to produce a context vector **z**_*t*_. This mechanism identifies the visits and clinical variables most relevant to the phenotype assignment. A trajectory-aware contrastive objective further organizes the context space by encouraging histories with similar outcomes to remain close while preserving distinctions in their longitudinal progression patterns. As a result, patients with the same diagnosis but different clinical trajectories can be assigned to different phenotypes, reducing the collapse of distinct progression pathways into a single group. The attention weights provide phenotype-level saliency by attributing each assignment to specific variables at specific visits.

### Phenotype bottleneck and prediction (§B.4)

The context vector **z**_*t*_ is assigned to *K* predictive phenotypes using a Dirichlet parameterization with concentration parameters ***ϕ***_*t*_ = *f*_sel_(**z**_*t*_) produced by the phenotype selector, where *f*_sel_ denotes the phenotype selector and each *ϕ*_*t,k*_ controls the strength of assignment toward phenotype *k*. During training, the soft phenotype assignment is sampled as ***π***_*t*_ ~ Dir(***ϕ***_*t*_), while at inference it is replaced by the Dirichlet mean 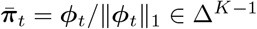. Each predictive phenotype *k* is associated with a learned phenotype embedding ***ϵ***_*k*_, and the patient-specific phenotype representation is given by the assignment-weighted mixture 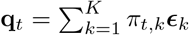. Together with the encoded static covariates **x**^stat^, **q**_*t*_ provides the prediction pathway from the longitudinal history to the downstream diagnosis **ŷ**_*t*+*h*_. When *K* > *C*, multiple predictive phenotypes can correspond to the same diagnostic outcome, enabling finer-grained progression phenotypes within a diagnostic class. A balancing regularizer prevents collapse of the phenotype space.

The training is done in three main stages to provide a stable initialization for predictive phenotype discovery before jointly optimizing the full architecture. Stage 1 trains the input embedding, encoders, and attention using the auxiliary diagnostic, feature-forecasting, trajectory-aware contrastive, and disentanglement objectives. Stage 2 initializes the phenotype embeddings from the resulting contexts and pretrains the selector and decoder using the selector and classification objectives, while keeping the upstream representation frozen to align the randomly initialized selector with an initial hard assignment. Stage 3 jointly optimizes the full architecture end-to-end using the final classification, contrastive, disentanglement, and phenotype-balancing objectives. Training schedules and implementation details are given in Appendix B.5. At inference, each patient’s window is passed through the learned encoders and dual attention to obtain the context **z**_*t*_, which is mapped by the predictive phenotype layer to a soft phenotype assignment 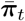 and then to the diagnosis prediction *ŷ*_*t*+*h*_. The corresponding attention weights provide visit–variable attributions for interpretability of predictions.

## 4 Experimental setup

The evaluation considers two complementary aspects: (i) the quality and interpretability of the discovered phenotype space, and (ii) downstream multi-horizon prediction. Dataset construction, preprocessing, implementation, hyperparameter search, and optimization procedures are in Appendix C.

Data used in the preparation of this article were obtained from the Alzheimer’s Disease Neuroimaging Initiative (ADNI) database (adni.loni.usc.edu). The ADNI was launched in 2003 as a public-private partnership, led by Principal Investigator Michael W. Weiner, MD. The primary goal of ADNI has been to test whether serial magnetic resonance imaging (MRI), positron emission tomography (PET), other biological markers, and clinical and neuropsychological assessment can be combined to measure theprogression of mild cognitive impairment (MCI) and early Alzheimer’s disease (AD) [12] C.1. Following prior works [5, 8, 13], each patient is represented by 21 variables (5 static and 16 time-varying) covering demographic, cognitive, clinical, and structural MRI information. After preprocessing, the dataset includes 1,788 participants and 10,941 visits. The task is three-class diagnosis prediction among Cognitively Normal (CN), Mild Cognitive Impairment (MCI), and Alzheimer’s Disease (AD) at the current visit (*h* = 0) and at *h* = 12 and *h* = 24 months using rolling longitudinal windows.

### Baselines

TRACE is compared with representative approaches for temporal clustering, interpretable temporal phenotyping, and longitudinal representation learning: KM-DTW [14, 15], AC-TPC, KM-E2P(**z**), KM-E2P(*y*) [5], T-Phenotype [8], and CAMELOT [6].

### Evaluation

All methods use the same patient-level 70%/10%/20% split with training-set standardization, and hyperparameters are selected with Optuna under the same search protocol and random seeds. Phenotype quality is evaluated with cluster Purity, Normalized Mutual Information (NMI) [16], Adjusted Rand Index (ARI) [17], and Silhouette Index (SI) [18], and prediction with Accuracy, F1, AUROC, and AUPRC. The diagnostic metrics are macro averaged.

## Results

Experiments are first performed in a label-matched setting, *K* = *C*, to enable fair comparison with prior methods, and then extended to *K* > *C*, enabling additional phenotype discovery.

### Label-matched phenotyping (*K* = *C*)

TRACE provides the strongest overall performance, achieving the best Purity, NMI, ARI, and SI as well as the highest Accuracy and macro-F1 across all prediction horizons (Tables 1 and 2). The dual attention further provides an interpretable view of the learned phenotype assignments through distinct visit–variable saliency patterns (Fig. 2). Its advantage becomes larger at longer horizons: at *h* = 24, TRACE improves over the strongest competing method by 3.1, 2.7, 3.5, and 5.8 percentage points in Purity, NMI, ARI, and SI, respectively, while significantly outperforming all baselines in Accuracy at both future horizons. T-Phenotype is the closest competitor, retaining the highest AUROC and AUPRC at *h* = 12 and *h* = 24, while KM-E2P(*y*) is strongest at *h* = 0 on AUROC and AUPRC. The strong performance of TRACE and T-Phenotype across horizons is consistent with their explicit use of longitudinal feature trajectories, which allows the learned phenotypes to reflect how clinical features evolve rather than only their current values. TRACE, however, constrains downstream prediction to operate through the predictive phenotype layer, making phenotype assignment directly responsible for the final prediction. While this can reduce the granularity of prediction scores, and consequently AUROC/AUPRC, relative to methods that retain richer continuous representations, it becomes advantageous at longer horizons: from *h* = 0 to *h* = 24, TRACE loses only 7.7 Accuracy points and 15.6 NMI points, compared with 9.9 and 16.5 for T-Phenotype, respectively. This slower degradation suggests that the TRACE phenotypes retain more progression-relevant information as the prediction horizon increases. KM-DTW, whose phenotype assignments are not produced on a supervised prediction path, remain far behind on phenotype quality at every horizon. Appendix C.2.2 explains why Purity and Accuracy coincide at *K* = *C* for the methods that predict through the phenotype assignment.

**Table 1:** Phenotype quality on the ADNI cohort across all prediction horizons. Values are mean ± standard deviation (%) over 10 random seeds. Bold marks the best and underline the second-best value in each column.

| Method | $h = 0$ months | | | | $h = 12$ months | | | | $h = 24$ months | | | |
| --- | --- | --- | --- | --- | --- | --- | --- | --- | --- | --- | --- | --- |
|  | Purity | NMI | ARI | SI | Purity | NMI | ARI | SI | Purity | NMI | ARI | SI |
| KM-DTW | 58.3±1.6* | 26.1±1.7* | 19.7±1.2* | 9.6±1.3* | 56.0±4.2* | 22.3±5.7* | 17.8±4.7* | 7.1±4.5* | 56.0±2.6* | 22.7±3.1* | 17.3±2.4* | 6.8±1.5* |
| AC-TPC | 84.9±1.2* | 55.1±2.4* | 58.3±3.1* | 57.8±3.1* | 77.2±2.7* | 43.1±3.8* | 42.1±5.1* | 39.5±6.1* | 71.3±3.3* | 35.1±4.2* | 33.0±5.6* | 27.7±6.4* |
| KM-E2P(z) | 80.3±1.3* | 48.9±2.6* | 48.4±2.8* | 46.4±3.2* | 75.4±2.8* | 43.6±3.9* | 40.1±4.9* | 35.5±5.9* | 72.0±4.4* | 36.8±6.3* | 34.5±7.5* | 29.1±8.6* |
| KM-E2P(y) | 86.0±0.9 | 57.1±2.0 | 60.9±2.4 | 60.5±2.4 | 78.3±2.2* | 43.8±3.6* | 43.8±4.4* | 41.9±4.9* | 73.6±3.6* | 36.6±5.7* | 35.8±5.7* | 32.0±7.6* |
| T-Phenotype | 85.6±1.2 | 56.9±2.2 | 61.7±3.1 | 60.3±2.9 | 78.9±1.8* | 45.5±3.1* | 45.4±4.0* | 43.2±4.3* | 75.4±4.7* | 40.4±6.3 | 40.1±8.0 | 36.1±9.3* |
| CAMELOT | 85.5±0.9* | 57.0±2.4 | 60.5±2.4 | 59.3±2.4* | <u>79.2±3.0*</u> | <u>46.3±5.0</u> | <u>46.6±6.2*</u> | <u>44.2±7.0*</u> | <u>75.7±3.2*</u> | <u>36.6±4.7*</u> | <u>40.3±6.0</u> | <u>36.2±6.9*</u> |
| TRACE | **86.5±1.4** | **58.7±2.8** | **62.4±3.3** | **62.0±3.7** | **81.6±1.3** | **49.4±2.6** | **50.8±3.0** | **50.5±3.2** | **78.8±2.2** | **43.1±3.9** | **43.8±4.8** | **42.0±5.0** |

**Table 2:** Downstream prediction performance on the ADNI cohort across all prediction horizons. Values are mean ± standard deviation (%) over 10 random seeds. Bold marks the best and underline the second-best value in each column.

| Method | $h = 0$ months | | | | $h = 12$ months | | | | $h = 24$ months | | | |
| --- | --- | --- | --- | --- | --- | --- | --- | --- | --- | --- | --- | --- |
|  | Acc | F1 | AUROC | AUPRC | Acc | F1 | AUROC | AUPRC | Acc | F1 | AUROC | AUPRC |
| KM-DTW† | 58.3±1.6* | 59.0±2.2* | 74.3±1.1* | 53.5±1.3* | 54.9±4.7* | 53.5±7.8* | 72.6±3.0* | 51.4±4.0* | 54.2±3.4* | 51.2±6.5* | 71.8±1.7* | 51.4±1.7* |
| AC-TPC | 84.9±1.2* | 85.0±1.1* | 83.3±14.0* | 75.1±16.6* | 77.2±2.7* | 77.8±2.5* | 83.5±10.8* | 75.3±11.8* | 71.3±3.3* | 71.9±3.0* | 84.2±4.2* | 73.9±6.5* |
| KM-E2P(z) | 80.3±1.3* | 80.5±1.2* | 93.4±1.2* | 87.6±2.3* | 75.4±2.8* | 76.2±2.5* | 91.6±1.9 | 84.9±2.9 | 72.0±4.4* | 72.4±4.1* | 88.1±3.2 | 79.3±4.8 |
| KM-E2P(y) | 86.0±0.9 | 86.0±0.9 | **96.1±0.4** | **92.8±0.7** | 78.3±2.2* | 78.7±2.2* | 92.3±1.2 | 86.0±2.3 | 73.6±3.4* | 73.9±3.4* | 89.0±2.4 | 80.8±3.8 |
| T-Phenotype | 85.6±1.2 | 85.6±1.0 | 94.5±0.5 | 92.5±0.8 | 78.9±1.8* | 79.3±1.8* | **92.8±0.9** | **86.9±1.5** | 75.7±4.4* | 76.0±4.2 | **90.3±2.9** | **83.0±4.5** |
| CAMELOT | 85.5±0.9* | 85.3±1.0* | 91.4±0.9* | 79.3±1.3* | <u>79.2±3.0*</u> | <u>79.7±2.8*</u> | 87.3±2.0* | 72.5±3.3* | <u>75.7±3.2*</u> | <u>75.8±3.0*</u> | 84.5±2.1* | 67.8±3.2* |
| TRACE | **86.5±1.4** | **86.5±1.3** | 94.3±0.9 | 89.5±2.2 | **81.6±1.3** | **82.2±1.4** | 92.7±0.6 | 85.6±1.7 | **78.8±2.2** | **78.5±2.3** | 89.3±2.1 | 80.0±4.5 |

\* Baseline significantly outperformed by TRACE ( $p < 0.05$ , one-sided Welch's $t$ -test over the 10 per-seed results.). The test is conducted separately for each baseline, metric, and horizon, with the alternative in the direction of TRACE's observed difference. † KM-DTW provides no predictive head. Its prediction metrics are obtained by assigning each phenotype its most frequent endpoint class and scoring each window's actual label against the average outcome of its assigned phenotype.

**Figure 2:**
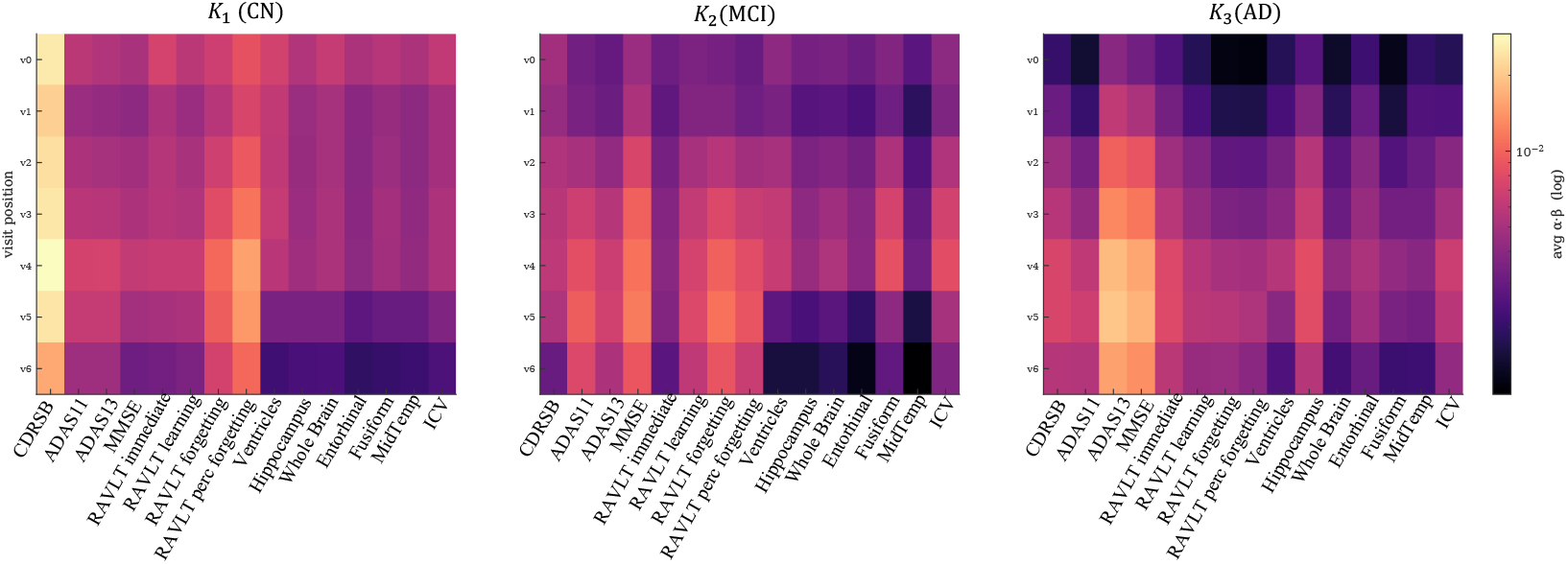
Saliency maps for the three phenotypes at *K* = *C* on ADNI. Each heatmap shows the average saliency score, *α β*, across the seven visits (*v*_0_–*v*_6_) and clinical variables. Brighter regions indicate features and visits that contribute more strongly to the phenotype assignment. The saliency patterns differ across phenotypes and evolve over the history window, with later visits and cognitive and memory measures becoming increasingly prominent for the MCI and AD phenotypes.

The accompanying visit–variable saliency 2 further explains the features and visits mostly contributing to the predictions for each phenotype for *h* = 24, and *K* = *C*. The maps show that TRACE assigns different importance patterns to different stages of the longitudinal history rather than relying uniformly on the same measurements. For the CN-dominant phenotype (*k*_0_), saliency is relatively concentrated on CDRSB and RAVLT-related measures, with relevant signals already present in the earlier visits and remaining comparatively localized throughout the sequence. The MCI-dominant phenotype (*k*_1_) exhibits a broader pattern of saliency across cognitive and memory variables, with stronger responses emerging in the middle and later visits, particularly for ADAS11/13, MMSE, and the RAVLT measures. For the AD-dominant phenotype (*k*_2_), saliency becomes most pronounced in the later visits and is concentrated primarily on cognitive and memory measures, including ADAS13, MMSE, and RAVLT variables. These patterns indicate that phenotype assignments depend on both *which* clinical features are informative and *when* those features become informative, providing an interpretable view of the longitudinal evidence used by TRACE to distinguish CN, MCI, and AD.

### Phenotype discovery beyond diagnostic labels (*K* > *C*)

Fixing *K* = *C* isolates differences in representation learning, but it assumes a one-to-one correspondence between diagnosis and phenotype and can obscure heterogeneity in how patients within the same diagnosis progress over time. TRACE is therefore evaluated with *K* > *C*, with *K* selected on the validation split from 4, …, 10 separately for each prediction horizon based on clustering coherence and alignment with the endpoint labels, allowing multiple latent phenotypes to emerge within the same outcome class C.4. Evaluated at *h* = 24 yields a fourth phenotype, referred to as *k*_3_, alongside the CN-, MCI-, and AD-dominant phenotypes *k*_0_, *k*_1_, and *k*_2_ obtained at *K* = *C*. Figure 3(a) compares the two partitions of the same evaluated windows and shows transitions of instances in phenotypes formed in the label-matched setting to the setting with *K* > *C*: the 87 instances in *k*_3_ are drawn from *k*_0_ and *k*_1_, and essentially none from the AD-dominant *k*_2_. The heterogeneity absorbed at *K* = *C* is therefore concentrated within the CN-and MCI-dominant groups. Figure 3(b) shows the corresponding mean feature trajectories over the seven window positions preceding the decision point. *k*_3_ is not simply an intermediate level of severity: on the cognitive and functional scales (CDRSB, ADAS13, RAVLT percent forgetting) it lies between *k*_0_ and *k*_1_ and stays closer to the CN-dominant trajectory, whereas its hippocampal volume trajectory is indistinguishable from the MCI-dominant *k*_1_ across the whole window. Appendix Fig. A1 reproduces this pattern in pairs of clinical variables for their evolution from the beginning to the end of the window, where *k*_3_ lies between *k*_0_ and *k*_1_. This cross-domain dissociation suggests a subgroup that a single diagnostic label does not capture, and supports the interpretation that TRACE does not merely split diagnostic labels arbitrarily: the additional phenotype captures a recognizable subset of trajectories within existing diagnostic classes.

**Figure 3:**
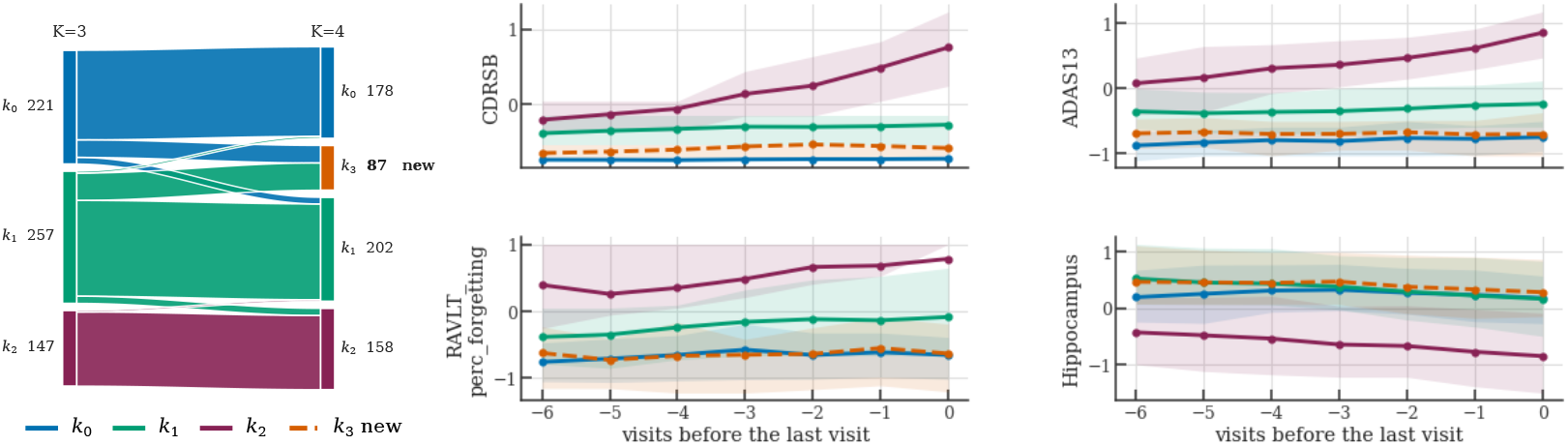
Phenotype discovery beyond diagnostic labels at *K* = 4 > *C* and *h* = 24 months. **(a)** Reassignment of the same evaluated windows between the label-matched (*K* = 3) and discovered (*K* = 4) partitions. Band widths give window counts, and the new phenotype *k*_3_ (87 windows) draws almost only from the CN-dominant *k*_0_ and the MCI-dominant *k*_1_. **(b)** Mean standardized feature trajectories per phenotype over the seven window positions preceding the decision point (position 0), shading spans one within-phenotype standard deviation.

## 6 Conclusion

A patient’s trajectory is most informative when read against others following a similar course rather than against the population as a whole. TRACE provides such a reference frame by learning outcome-predictive progression phenotypes that capture both clinical state and trajectory-specific structure. On ADNI, TRACE improves over existing temporal phenotyping methods at every horizon, with the largest margins at 24 months. In the K>C setting, TRACE reveals progression heterogeneity that is obscured by diagnostic labels, identifying a phenotype between CN- and MCI-dominant trajectories with distinct cross-domain patterns. TRACE’s visit- and feature-level attention characterizes the observations underlying each phenotype, highlighting which clinical features and visits warrant closer longitudinal attention. Together, outcome-predictive phenotype discovery and interpretable trajectory characterization provide a finer-grained view of progression than diagnostic labels alone and may support more timely clinical reassessment when a patient’s trajectory begins to diverge from its current diagnostic category. These findings demonstrate the potential of TRACE to uncover progression structure that is invisible when patients are organized only by diagnostic labels. However, the current evaluation is limited to ADNI, where Alzheimer’s disease progresses slowly and seven available visits span only approximately 3.5 years, providing limited observable temporal heterogeneity. The discovered phenotypes remain exploratory and require external replication, stability analysis, and independent clinical validation, particularly when *K* > *C*, where diagnosis-based metrics provide limited evidence of phenotype quality. Further component ablations and quantitative assessments of attention faithfulness are also needed to establish which modeling choices drive the observed gains and how reliably the explanations reflect the model’s behavior. Evaluation on cohorts with denser measurements and more heterogeneous progression patterns is needed to establish the temporal modeling capabilities of TRACE, as well as their utility for longitudinal clinical monitoring. Future work will also explicitly investigate transitions between discovered phenotypes to determine whether they correspond to clinically meaningful progression pathways.

## Data Availability

The data used in in this study is from Alzheimer's Disease Neuroimaging Initiative (ADNI). To access the data, please follow instructions provided by ADNI.

https://adni.loni.usc.edu/

## Acknowledgments

This research was partly supported by NSF Grant 2112533.

The experiments are performed using the dataset from Alzheimer’s Disease Neuroimaging Initiative (ADNI). Data collection and sharing for the ADNI is funded by the National Institute on Aging (National Institutes of Health Grant U19 AG024904). The grantee organization is the Northern California Institute for Research and Education. In the past, ADNI has also received funding from the National Institute of Biomedical Imaging and Bioengineering, the Canadian Institutes of Health Research, and private sector contributions through the Foundation for the National Institutes of Health (FNIH) including generous contributions from the following: AbbVie, Alzheimer’s Association; Alzheimer’s Drug Discovery Foundation; Araclon Biotech; BioClinica, Inc.; Biogen; Bristol-Myers Squibb Company; CereSpir, Inc.; Cogstate; Eisai Inc.; Elan Pharmaceuticals, Inc.; Eli Lilly and Company; EuroImmun; F. Hoffmann-La Roche Ltd and its affiliated company Genentech, Inc.; Fujirebio; GE Healthcare; IXICO Ltd.; Janssen Alzheimer Immunotherapy Research & Development, LLC.; Johnson & Johnson Pharmaceutical Research &Development LLC.; Lumosity; Lundbeck; Merck & Co., Inc.; Meso Scale Diagnostics, LLC.; NeuroRx Research; Neurotrack Technologies; Novartis Pharmaceuticals Corporation; Pfizer Inc.; Piramal Imaging; Servier; Takeda Pharmaceutical Company; and Transition Therapeutics.

## A APPENDIX

The appendix specifies TRACE from input construction to training and evaluation. Section B presents the forward computation in data-flow order, from input representation through temporal encoding, phenotype discovery, and prediction. Section C specifies the cohort, preprocessing, evaluation metrics, the hyperparameter search space, and the criteria used to select the number of phenotypes.Table A1 collects the notation used throughout the paper.

**Table A1:** Notation used throughout the paper. Parameters selected by validation-set search, including loss weights, optimization and schedule parameters, and searched architecture widths, are reported only in Section C.3.

| Symbol | Meaning | Shape / Domain |
| --- | --- | --- |
| *Data and indexing* |  |  |
| $N, N_W$ | Number of patients and evaluated windows | $\mathbb{N}$ |
| $W$ | Window length | $\mathbb{N}$ |
| $F, S$ | Longitudinal and static input channels | $\mathbb{N}$ |
| $C, K$ | Outcome classes and phenotype components | $\mathbb{N}$ |
| $i, t$ | Visit index and decision-point position within a window | $\{1, \dots, W\}$ |
| $w$ | Window index; $a, b$ index the two windows of a contrastive pair | — |
| $f, c, k$ | Channel, class, and phenotype indices | — |
| $h, \rho$ | Prediction horizon and forecast-target acceptance band | — |
| *Window inputs and masks* |  |  |
| $\mathbf{x}_i, \mathbf{m}_i$ | Longitudinal features and missingness mask at visit $i$ | $\mathbb{R}^F, \{0, 1\}^F$ |
| $\tau_i, \delta_i$ | Absolute visit time and inter-visit time $\tau_i - \tau_{i-1}$ | $\mathbb{R}$ |
| $\mathcal{V}$ | Valid visit positions | $\{1, \dots, W\}$ |
| $\mathcal{B}$ | Mini-batch of windows | — |
| $\mathcal{P}_k, \mathcal{Y}_c$ | Windows assigned to phenotype $k$ and windows of class $c$ | — |
| *Representations* |  |  |
| $d, d_z$ | Encoder and phenotype-path widths | $\mathbb{N}$ |
| $d_\tau, d_e, d_s$ | Time, channel, and static embedding widths | 16 |
| $\mathbf{e}_i$ | Visit embedding | $\mathbb{R}^d$ |
| $\mathbf{h}_i^d, \mathbf{h}_i^e$ | Diagnostic and evolution states | $\mathbb{R}^d$ |
| $\mathbf{h}_i^f$ | Concatenated encoder state | $\mathbb{R}^{2d}$ |
| $\mathbf{x}^{\text{stat}}$ | Static covariates, encoded to $\mathbb{R}^{d_s}$ at the decoder | $\mathbb{R}^S$ |
| $\ell_i$ | Temporal-attention logit at visit $i$ | $\mathbb{R}$ |
| $\alpha_{t,i}$ | Visit-level attention weight | $[0, 1]$ |
| $\beta_i$ | Channel-level attention weights | $\Delta^{F-1}$ |
| $\mathbf{z}_t, \mathbf{W}_z$ | Context of the longitudinal window and its output projection | $\mathbb{R}^{d_z}, \mathbb{R}^{d_z \times F}$ |
| *Phenotypes and prediction* |  |  |
| $k_t, k_w$ | Latent phenotype at the decision point, hard assignment of window $w$ | $\{1, \dots, K\}$ |
| $\tilde{k}_t$ | Initial hard phenotype assignment | $\{1, \dots, K\}$ |
| $\phi_t$ | Dirichlet concentration vector | $[1, \phi_{\max}]^K$ |
| $\pi_t, \bar{\pi}_t$ | Sampled and mean soft phenotype assignments | $\Delta^{K-1}$ |
| $\epsilon_k$ | Learned embedding of predictive phenotype $k$ | $\mathbb{R}^{d_z}$ |
| $\mathbf{q}_t$ | Phenotype representation (mixture of $\epsilon_k$) | $\mathbb{R}^{d_z}$ |
| $\hat{\mathbf{y}}_{t+h}$ | Predicted outcome distribution | $\Delta^{C-1}$ |
| *Trajectory-aware contrastive objective* |  |  |
| $s_{ab}^{\text{out}}, s_{ab}^{\text{traj}}$ | Outcome and trajectory similarity of pair $(a, b)$ | $\{0, 1\}, [0, 1]$ |
| $w_{ab}^{\text{att}}, w_{ab}^{\text{rep}}, \Lambda_{ab}$ | Attraction, repulsion, and total pair weight | $\mathbb{R}_{\geq 0}$ |
| $\mu_{\text{att}}, \mu_{\text{rep}}$ | Attraction and repulsion parameters | $[0, 1]$ |
| $\nu_{\text{att}}, \nu_{\text{rep}}$ | Cross-quadrant attraction and repulsion parameters | $[0, 1]$ |
| $\kappa, Z_a$ | Softmax temperature and anchor attraction mass | $\mathbb{R}_{>0}$ |
| $q_1, \dots, q_4$ | Equilibrium ratios of the four outcome–trajectory quadrants | $[0, 1]$ |

### B Method Details

This section details the TRACE architecture and training objectives. Tables A2 and A3 provide a compact summary of the modules and training objectives, respectively.

**Table A2:** Module-level specification of TRACE, in forward order. Superscripts denote the training stages in which a loss is active. Within each row, **bold losses directly supervise the module**, whereas regular losses provide indirect supervision through downstream components.

| Module | Input → Output | Supervision | Role |
| --- | --- | --- | --- |
| **Input representation** (Section B.1) |  |  |  |
| Numerical embedding<br>$\mathbf{g}_f(\cdot, \cdot)$ | $x_{i,f}, m_{i,f} \rightarrow \tilde{\mathbf{g}}_{i,f}$ | $\mathcal{L}^{(1)}, \mathcal{L}^{(3)}$ | Variable-specific encoding with explicit missingness. |
| Visit projection<br>$\mathbf{W}^{\text{proj}}$ | $\tilde{\mathbf{g}}_{i,\cdot} \rightarrow \mathbf{e}_i^{\text{ch}}$ | $\mathcal{L}^{(1)}, \mathcal{L}^{(3)}$ | Mixes the $F$ channels into a visit representation. |
| Time2Vec | $\tilde{\tau}_i, \tilde{\delta}_i \rightarrow \mathbf{e}_i^\tau$ | $\mathcal{L}^{(1)}, \mathcal{L}^{(3)}$ | Encodes absolute time and inter-visit gaps. |
| Visit embedding | $\mathbf{e}_i^{\text{ch}}, \mathbf{e}_i^\tau \rightarrow \mathbf{e}_i = [\mathbf{e}_i^{\text{ch}}; \mathbf{e}_i^\tau]$ | — | Parameter-free concatenation, in $\mathbb{R}^d$; the sequence $\mathbf{e}_{1:t}$ feeds both encoders. |
| **Dual-objective encoders** (Section B.2) |  |  |  |
| Diagnostic encoder<br>$\text{Enc}^d(\cdot)$ | $\mathbf{e}_{1:t} \rightarrow \mathbf{h}_{1:t}^d$ | $\mathcal{L}_{\text{aux}}^{(1)}, \mathcal{L}_{\text{dis}}^{(1,3)}, \mathcal{L}_{4q}^{(1,3)}, \mathcal{L}_{\text{cls}}^{(3)}, \mathcal{L}_{\text{bal}}^{(3)}$ | Encodes outcome-relevant diagnostic state. |
| Evolution encoder<br>$\text{Enc}^e(\cdot)$ | $\mathbf{e}_{1:t} \rightarrow \mathbf{h}_{1:t}^e$ | $\mathcal{L}_{\text{fcst}}^{(1,3)}, \mathcal{L}_{\text{dis}}^{(1,3)}, \mathcal{L}_{4q}^{(1,3)}, \mathcal{L}_{\text{cls}}^{(3)}, \mathcal{L}_{\text{bal}}^{(3)}$ | Encodes trajectory dynamics and future measurement trends. |
| Auxiliary head<br>$\mathbf{W}^{\text{aux}}$ | $\mathbf{h}_t^d \rightarrow \hat{\mathbf{y}}_{t+h}^{\text{aux}}$ | $\mathcal{L}_{\text{aux}}^{(1)}$ | Temporary endpoint-prediction head for diagnostic pretraining. |
| Forecasting head<br>$\mathbf{W}^{\text{fc}}$ | $\mathbf{h}_t^e \rightarrow \hat{\mathbf{x}}_{t+h}$ | $\mathcal{L}_{\text{fcst}}^{(1,3)}$ | Temporary measurement-prediction head for trajectory supervision. |
| **Dual attention and context** (Section B.3) |  |  |  |
| Stream fusion | $\mathbf{h}_i^d, \mathbf{h}_i^e \rightarrow \mathbf{h}_i^f = [\mathbf{h}_i^d; \mathbf{h}_i^e]$ | — | Parameter-free concatenation, in $\mathbb{R}^{2d}$. Both attentions read diagnostic state and trajectory jointly. |
| Temporal attention<br>$\mathbf{W}_\alpha$ | $\mathbf{h}_{1:t}^f \rightarrow \{\alpha_{t,i}\}_{i \leq t}$ | $\mathcal{L}_{4q}^{(1,3)}, \mathcal{L}_{\text{cls}}^{(3)}, \mathcal{L}_{\text{bal}}^{(3)}$ | One weight per visit, with $\sum_{i \leq t} \alpha_{t,i} = 1$: how much visit $i$ counts towards the decision at $t$. |
| Variable attention<br>$\mathbf{W}_\beta$ | $\mathbf{h}_i^f, \mathbf{m}_i \rightarrow \beta_i$ | $\mathcal{L}_{4q}^{(1,3)}, \mathcal{L}_{\text{cls}}^{(3)}, \mathcal{L}_{\text{bal}}^{(3)}$ | One weight per variable within visit $i$. The mask $\mathbf{m}_i$ drops unobserved entries from the softmax. |
| Context projection<br>$\mathbf{W}_z$ | $\sum_{i \leq t} \alpha_{t,i} (\beta_i \odot \mathbf{x}_i) \rightarrow \mathbf{z}_t$ | $\mathcal{L}_{4q}^{(1,3)}, \mathcal{L}_{\text{cls}}^{(3)}, \mathcal{L}_{\text{bal}}^{(3)}$ | Both attentions are applied to the raw measurements, so the summary stays in clinical-variable units before projection to $\mathbb{R}^{dz}$. |
| **Predictive phenotype layer** (Section B.4) |  |  |  |
| Selector MLP<sub>sel</sub>(·) | $\mathbf{z}_t \rightarrow \phi_t, \pi_t$ | $\mathcal{L}_{\text{sel}}^{(2)}, \mathcal{L}_{\text{cls}}^{(3)}, \mathcal{L}_{\text{bal}}^{(3)}$ | Produces soft phenotype assignments. |
| Phenotype embeddings $\epsilon_k$ | $\pi_t \rightarrow \mathbf{q}_t$ | $\mathcal{L}_{\text{cls}}^{(3)}$ | Define the learnable phenotype anchors. |
| Static encoder<br>$\text{Enc}^{\text{stat}}(\cdot)$ | $\mathbf{x}^{\text{stat}} \rightarrow \mathbf{s}$ | $\mathcal{L}_{\text{cls}}^{(2,3)}$ | Encodes static covariates for outcome decoding. |
| Decoder<br>$\text{MLP}_{\text{dec}}(\cdot)$ | $\mathbf{q}_t, \mathbf{s} \rightarrow \hat{\mathbf{y}}_{t+h}$ | $\mathcal{L}_{\text{cls}}^{(2,3)}$ | Maps phenotype context to the outcome distribution. |

**Table A3:** Training objectives in TRACE across the three training stages(Section B.5). Stage 1 and Stage 3 optimize the weighted objectives *L*^1^ (Eq. (16)) and *L*^3^ (Eq. (18)), respectively, while Stage 2 applies its two objectives sequentially.

| Loss | Target / comparison | Stage | Updated parameters | Role |
| --- | --- | --- | --- | --- |
| $\mathcal{L}_{\text{aux}}$ | $\hat{\mathbf{y}}_{t+h}^{\text{aux}}$ vs. endpoint label $\mathbf{y}_{t+h}$ | 1 | Auxiliary head, $\text{Enc}^d$, input embedding | Establishes the diagnostic stream as outcome-relevant before introducing the phenotype layer. |
| $\mathcal{L}_{\text{fcast}}$ | $\hat{\mathbf{x}}_{t+h}$ vs. measurements at the visit nearest $\tau_t + h$ | 1, 3 | Forecasting head, $\text{Enc}^e$, input embedding | Encourages the evolution stream to encode information predictive of future measurements. |
| $\mathcal{L}_{\text{dis}}$ | $\mathbf{h}_w^d$ vs. $\mathbf{h}_w^e$ through a CLUB mutual-information bound | 1, 3 | Both encoders, input embedding, $\xi$ via a separate optimizer | Discourages redundant information across the diagnostic and evolution streams. |
| $\mathcal{L}_{4q}$ | $\cos(\mathbf{z}_a, \mathbf{z}_b)$ vs. outcome agreement $s_{ab}^{\text{out}}$, weighted by trajectory similarity $s_{ab}^{\text{traj}}$ | 1, 3 | $\mathbf{W}_z$, attention, both encoders, input embedding | Separates outcome agreement from trajectory similarity, with outcome determining pairwise direction and trajectory determining its strength. |
| $\mathcal{L}_{\text{sel}}$ | $\pi_t$ vs. pseudo-label $\tilde{k}_t$ from Eq. (17) | 2 | Selector only | Initializes the selector to recover the prototype-based partition. |
| $\mathcal{L}_{\text{cls}}$ | $\hat{\mathbf{y}}_{t+h}$ decoded from sampled $\pi_t$ vs. $\mathbf{y}_{t+h}$ | 2, 3 | Decoder and $\text{Enc}^{\text{stat}}$ in Stage 2 and $\epsilon_k$, selector, and upstream modules in Stage 3 | Provides endpoint supervision for learning the phenotype embeddings and decoder. |
| $\mathcal{L}_{\text{bal}}$ | Batch occupancy $\bar{\pi}^B$ vs. $\text{Unif}(K)$ | 3 | Selector and upstream modules, excluding $\epsilon_k$ and decoder | Prevents phenotype collapse by encouraging all $K$ assignments to remain active. |

#### B.1 Window Construction and Input Embedding

Each training instance is one longitudinal window: the history *H*_*t*_ available at a decision point *t*, restricted to a fixed window length of *W* visits and left-padded when fewer than *W* visits are available, so that the decision point always occupies the last window position. Positions within a window are indexed by *i, V* and denotes the positions that carry a real visit. A window therefore supplies the measurements **x**_*i*_ ∈ ℝ^*F*^ of the *F* longitudinal channels, the missingness masks **m**_*i*_ ∈ {0, 1}^*F*^ with *m*_*i,f*_ = 1 when channel *f* is not measured at visit *i*, and the visit times *τ*_*i*_, together with the patient’s *S* static covariates **x**^stat^ ∈ ℝ^*S*^. Missing longitudinal values are imputed upstream (Section C.1) and remain in **x**_*i*_, but **m**_*i*_ records which entries were originally measured. All baselines receive the same imputed numerical input, whereas TRACE additionally receives **m**_*i*_ throughout its representation and prediction pipeline, allowing it to distinguish observed from imputed values without further imputation.

Static covariates are extracted before window construction, taking each variable from the first visit at which it is observed and setting it to zero when it is never observed. They are removed from the temporal stream, so that time-invariant variables cannot contribute to visit-or channel-level attention, and are reintroduced only at the decoder, through a static encoder 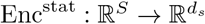, a small MLP of output width *d*_*s*_. All baselines retain the original channel representation.

Every term of the training objective is evaluated once per window, at its decision point, which is also the position read at inference. Each longitudinal channel is embedded independently into a *d*_*e*_-dimensional representation before any mixing across channels, allowing TRACE to capture channel-specific numerical structure. Following the numerical embedding families of Gorishniy et al. [19], three variants are considered: L applies a channel-specific linear projection 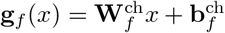 with 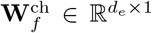 and 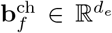. LR adds a ReLU nonlinearity. PLR first maps the scalar through *n*_*ω*_ learnable sinusoidal features, whose frequencies are channel-specific and initialized from *N*(0, 0.1^2^), then projects the resulting 2*n*_*ω*_-dimensional representation through a channel-specific 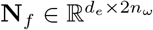. When a cell is unobserved, that is *m*_*i,f*_ = 1, its numerical embedding is replaced by a learned channel-specific missingness token 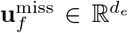, so that an imputed value is never embedded as if it had been measured.

The resulting channel embeddings 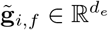 are concatenated across the *F* longitudinal variables and projected into the channel component of the visit representation, 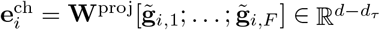 with 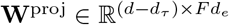, where *d* is the model width shared by the visit embedding and the two encoders and *d*_*τ*_ the number of its dimensions reserved for time. As a control, the none variant bypasses both the channel-wise embedding and the learned missingness tokens and instead projects the concatenation of the masked values and the mask itself, 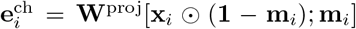.

Temporal information is encoded separately from the longitudinal measurements using two Time2Vec mappings [20], one for the absolute visit time *τ*_*i*_ and one for the inter-visit time *δ*_*i*_ = *τ*_*i*_ −*τ*_*i−* 1_. For an input scalar *u*, Time2Vec is t2v(*u*) = [*ω*_0_*u* + *φ*_0_; sin(***ω****u* + ***φ***)], with the linear and periodic coefficients learned. Both inputs are normalized using valid positions only, so that padding cannot affect the temporal scale: 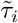 divides *τ*_*i*_ by the largest visit time among the positions in *V*, and 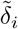 divides *δ*_*i*_ by their mean inter-visit time, each denominator floored away from zero. The two mappings contribute *d*_*τ*_ dimensions in total, split as evenly as possible between absolute and inter-visit time, and are concatenated with the channel representation to form the visit embedding

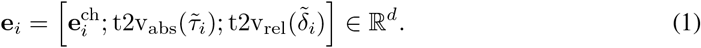

The resulting sequence **e**_1:*t*_ is passed to the diagnostic and evolution encoders described next.

#### B.2 Dual-Objective Longitudinal Representation Learning

TRACE learns two complementary causal representations of the same longitudinal history. The encoders Enc^*d*^ and Enc^*e*^ map the visit sequence to diagnostic and evolution states, 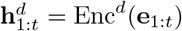 and 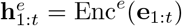 with 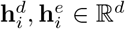, where each state depends only on positions up to *i*. The two encoders share the input representation but have separate parameters, allowing them to extract different temporal structure from the same measurements. Each encoder is a unidirectional LSTM, a GRU, or a causal Transformer with *n*_lay_ layers. The Transformer applies a causal self-attention mask, while the recurrent architectures are causal by construction. Dropout with rate *p*_drop_ is applied between layers, and padded positions are excluded throughout.

The two representations are given distinct predictive roles at the decision point: 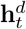 predicts the outcome at the future horizon 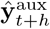, while 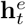 predicts the longitudinal measurements at the same future time 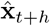. The diagnostic stream is trained by cross-entropy against the endpoint label **y**_*t*+*h*_, and the evolution stream by squared error against the measurements **x**_*t*+*h*_ recorded at the same future time,

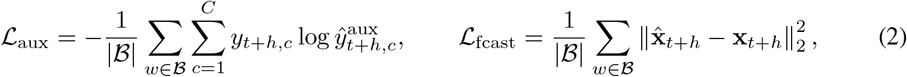

where *t* is the decision point of window *w* and **y**_*t*+*h*_ is the one-hot endpoint label. The two losses are computed only for the windows whose horizon-*h* target is observed, and the squared error is further restricted to the channels actually measured at that target visit. The forecasting target is resolved on the patient’s own clock rather than by a fixed index shift: it is read at the observed visit closest to *τ*_*t*_ + *h*, and a window contributes no forecasting term when no visit falls within the acceptance band *ρ* around that time. This is what keeps the two supervisions on one clock, since a fixed duration spans a patient- and visit-dependent number of positions. At *h* = 0 the label remains a genuine prediction, and the feature values at the decision point are reconstructed.

The two predictive objectives alone do not ensure that the streams capture distinct information. TRACE therefore adds a CLUB-based disentanglement objective [21] that minimizes an upper bound on the mutual information between the diagnostic and evolution representations. Intuitively, it compares how well the evolution representation can be predicted from its corresponding diagnostic representation against an evolution representation randomly paired from the same mini-batch. Specifically, a probabilistic model *q*_*ξ*_(**h**^*e*^ | **h**^*d*^) is learned to estimate the conditional distribution of the evolution representation given the diagnostic representation. TRACE computes the log-likelihood of the matched pair 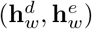 and subtracts the log-likelihood of a mismatched pair 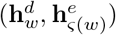, where *ς* randomly permutes the evolution representations within the mini-batch. The resulting CLUB objective is

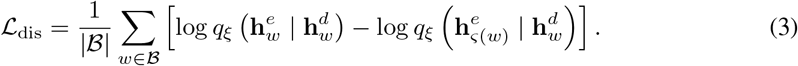

Here, the matched pairs retain the dependence between the two streams, whereas the random permutation breaks this correspondence and approximates independent samples. Minimizing their log-likelihood difference therefore discourages information in one stream from being predictable from the other, encouraging the diagnostic and evolution encoders to retain complementary information.

Together, *L*_aux_ and *L*_fcast_ define the complementary predictive roles of the two streams, while *L*_dis_ explicitly separates them. This separation is what the subsequent phenotype and trajectory objectives rely on: the diagnostic representation provides an outcome-oriented axis, whereas the evolution representation provides the trajectory-oriented axis. In the shared-encoder ablation, Enc^*d*^ and Enc^*e*^ are tied to a single set of parameters, so the two representations coincide, *L*_dis_ is identically zero, and it is disabled. The diagnostic and forecasting heads are used only during training and are discarded at inference, leaving the predictive phenotype layer as the sole path to the final outcome prediction.

#### B.3 Dual attention and saliency attribution

TRACE uses the dual-attention mechanism of TA-RNN [7] to form a context representation from the longitudinal trajectory. Temporal attention assigns a weight *α*_*t,i*_ to each visit,

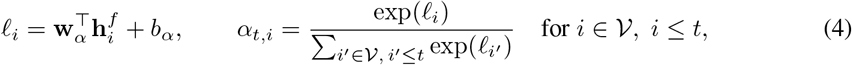

and *α*_*t,i*_ = 0 at padded positions, where 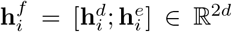 is the fused encoder state and **w**_*α*_ ∈ ℝ^2*d*^, *b*_*α*_ ∈ ℝ are learned. Variable attention assigns a weight *β*_*i,f*_ to each clinical variable within visit *i*,

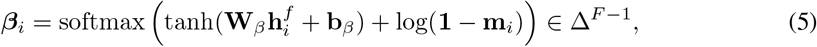

with learned **W**_*β*_ ∈ ℝ^*F×*2*d*^ and **b**_*β*_ ∈ ℝ^*F*^, where log 0 = −∞ removes the unobserved channels from the softmax. The missingness mask is applied *after* the tanh, since tanh(−∞) = −1 is a finite logit that would not exclude the channel. A position at which no channel is observed leaves the softmax undefined and is assigned ***β***_*i*_ = **0**, which removes it from the context. The resulting context is

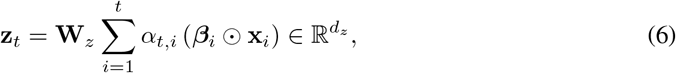

where 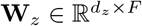 maps the attended measurements to the phenotype-path width *d*_*z*_.

##### Attention over raw measurements

Both attention maps are computed from the fused state 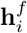, which carries the history up to visit *i*, but they are applied to the raw measurement vector **x**_*i*_ rather than to 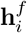 itself, and the only mixing across variables occurs in the final projection **W**_*z*_. The context of Eq. (6) is therefore a weighted combination of the recorded clinical variables in their own training-standardized units, which is what allows the attribution below to be read at the level of the data rather than of the representation: *β*_*i,f*_ is the share of visit *i* carried by variable *f*, so every attributed cell names an actual clinical variable together with the value it took at that visit. Attending over 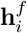 instead would place the weights on latent dimensions that are not identified with clinical variables, and variable-level importance would then have to be recovered by a post-hoc attribution method applied through the encoder, introducing an approximation between the explanation and the computation the model actually performs. Because the missingness mask in Eq. (5) removes unobserved entries from the softmax, attributed mass is assigned only to values that were measured, so the forward-filled and median-imputed entries of Section C.1 cannot be presented as evidence for a phenotype. The cost of this parameterization is that the context depends on the measurements linearly, with all nonlinearity confined to the weights *α*_*t,i*_ and ***β***_*i*_ and to the encoders that produce them, so the capacity of the model sits in what the attention selects rather than in how the selected values are combined.

##### Saliency attribution

Because the model makes an endpoint prediction from the history available at *t*, the two attention maps can be combined with the assignment 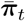 over the *K* phenotypes of Section B.4 into an endpoint-specific attribution tensor, one per window,

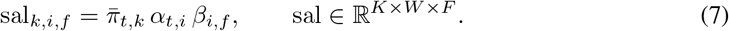

Since 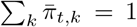, ∑_*i*_ *α*_*t,i*_ = 1, and ∑_*f*_ *β*_*i,f*_ = 1, the tensor sums to one, so sal_*k,i,f*_ is the fraction of the endpoint decision carried by variable *f* at visit *i* under phenotype *k*, and Eq. (7) is a decomposition of how the endpoint context was formed.

##### Trajectory-aware contrastive objective

The trajectory-aware contrastive objective is applied to every ordered pair (*a, b*) of distinct windows in the mini-batch and operates on their context representations **z**_*a*_.The resulting context space is therefore the same space used for phenotype clustering during *K*-selection and for phenotype assignment at inference (Section C.4). Each pair is placed by two similarities, both read at the decision point: outcome similarity is the agreement of the endpoint labels used by the classification loss, while trajectory similarity is the rescaled cosine similarity between the evolution states,

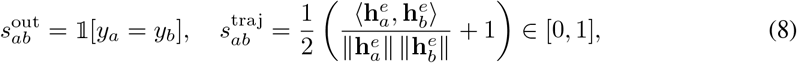

where *y*_*a*_ abbreviates the endpoint label of window *a*, and 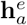 is detached from the computational graph so that the objective cannot reshape the axis it measures. An alternative trajectory measure, the displacement of **h**^*e*^ from the window’s first valid state, is also implemented but is held fixed rather than searched (Section C.3). The attraction and repulsion weights are

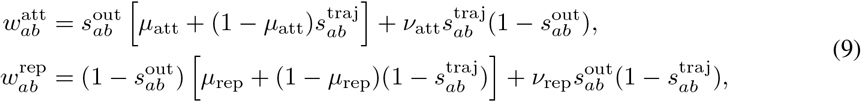

where *µ*_att_, *µ*_rep_ ∈ [0, 1] are the baseline attraction and repulsion applied to same- and different-outcome pairs irrespective of trajectory, *v*_att_, *v*_rep_ ∈ [0, 1] are the cross-quadrant weights that attract different-outcome pairs with similar trajectories and repel same-outcome pairs with dissimilar ones, and 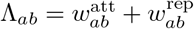 is the pair’s total weight. Thus, endpoint outcome determines the direction of the interaction, while trajectory similarity determines how strongly that interaction is applied. The contrastive objective, averaged over anchors, is

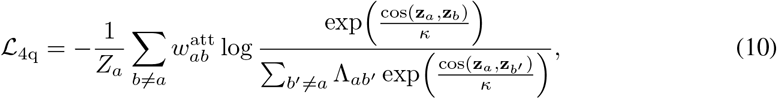

where *b* ranges over the other windows of the mini-batch, *κ* is the softmax temperature, and 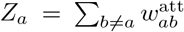 is anchor *a*’s total attraction mass. Setting the per-pair gradient of Eq. (10) to zero drives the weighted softmax mass on pair (*a, b*) to 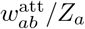, which holds exactly when 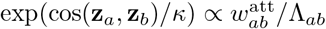 across the row. Evaluated at the extremes 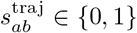, the four outcome–trajectory quadrants therefore have equilibrium ratios

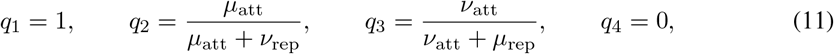

for same-outcome/similar-trajectory, same-outcome/dissimilar-trajectory, different-outcome/similar-trajectory, and different-outcome/dissimilar-trajectory pairs, respectively. The constraint *q*_3_ *< q*_2_, verified at construction, ensures that outcome agreement remains more influential than trajectory similarity. Eq. (11) also locates the objective within the contrastive family. At *µ*_att_ = *µ*_rep_ = 1 and *v*_att_ = *v*_rep_ = 0 the two mixed targets collapse onto the extremes, *q*_2_ = *q*_1_ = 1 and *q*_3_ = *q*_4_ = 0. The weights then reduce to 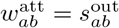 and 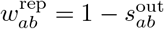, so Λ_*ab*_ ≡ 1, the trajectory similarity leaves the objective entirely, and *Z*_*a*_ counts the same-outcome partners of anchor *a*. Eq. (10) is then exactly the supervised contrastive loss of [22] in its 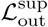 form — positives averaged outside the logarithm, which is the stronger of their two forms — averaged rather than summed over anchors. Restricting further to one positive per anchor, drawn by instance identity rather than by a shared label, leaves InfoNCE [23].

#### B.4 Predictive phenotype layer and prediction

The predictive phenotype layer, the phenotype bottleneck of Section 3, converts each window’s context **z**_*t*_ into a distribution over *K* predictive phenotypes, and keeps the assignment probabilistic through a Dirichlet selector [24] so that phenotype preference and confidence remain distinct. Because the diagnostic and forecasting heads of Section B.2 are discarded after training, this layer is the sole path from the observed history to the endpoint prediction, so the phenotype assignment is part of the forward predictive path rather than a post-hoc clustering of an already-trained representation. The selector maps the context to Dirichlet concentration parameters,

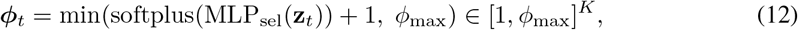

where 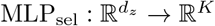 is a two-layer network. The lower bound and upper clamp serve different purposes: adding one ensures *ϕ*_*t,k*_ ≥ 1, giving a unimodal Dirichlet and preventing the density from concentrating at the simplex boundary before the objectives have shaped the assignment, while *ϕ*_max_ prevents unbounded concentration that would collapse the reparameterized sample toward a one-hot vector and cause numerical underflow in the Dirichlet entropy. The direction of ***ϕ***_*t*_ determines which phenotypes a window favors, while its total concentration ∥***ϕ***_*t*_∥ _1_ controls how strongly it favors them. Training therefore uses a reparameterized Dirichlet sample, whereas evaluation uses its mean,

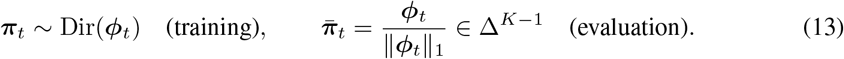

Sampling regularizes the phenotype layer by requiring the endpoint prediction to remain stable under perturbations of the assignment. The evaluation mean 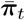 is the assignment used downstream for the phenotype factor of the saliency decomposition in Eq. (7), for the reported prediction path, and for the hardened partition evaluated by the clustering metrics of Section C.2. Each phenotype is represented by a learned phenotype embedding 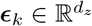, a representative vector of predictive phenotype *k* in the context space, and the phenotype representation **q**_*t*_ is the mixture of these embeddings weighted by the window’s phenotype assignment, which is then fused with the encoded static covariates for endpoint prediction,

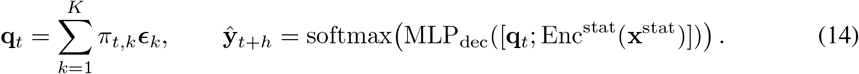

where MLP_dec_ is the decoder, a two-layer network of width *d*_dec_. Because ***π***_*t*_ lies on the simplex, **q**_*t*_ is a convex combination of the phenotype embeddings, so the context reaches the prediction only through the phenotype assignment. The same decoder is used during training and inference, differing only in whether it receives ***π***_*t*_ or 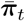. The predictive phenotype layer is shaped by three objectives applied in staged order. *L*_sel_ first aligns the selector with the initial phenotype-embedding partition while the upstream representation is frozen (Section B.4.1). *L*_cls_ then trains the phenotype layer for endpoint prediction and is the only objective that updates the phenotype embeddings and decoder (Section B.4.2), and *L*_bal_ prevents phenotype collapse during joint optimization (Section B.4.3). The choice of *K* is external to these objectives. When *K* > *C*, several predictive phenotypes may correspond to the same diagnostic outcome, which is what allows finer-grained progression phenotypes to be discovered.

##### B.4.1 Selector pretraining objective

In Stage 2, the selector is the only trainable module and is fitted to the pseudo-label 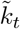 produced by the phenotype-embedding initialization of Eq. (17). The objective *L*_sel_ is the negative log-probability that the reparameterized sample places on that pseudo-label, log 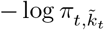, averaged over the mini-batch. The phenotype embeddings are held at their initialization, while the input embedding, encoders, attention module, and decoder remain frozen. This stage therefore gives the selector an initial correspondence between contexts and phenotype embeddings before the endpoint prediction objective is introduced.

##### B.4.2 Phenotype prediction objective

In the next stage, the previously frozen components are unfrozen and the model is trained jointly for endpoint prediction. The objective *L*_cls_ is the cross-entropy between the decoder output **ŷ**_*t*+*h*_ of Eq. (14) and the endpoint label, averaged over the mini-batch and, as for _aux_, computed only for the windows whose horizon-*h* label is observed. It is evaluated on the reparameterized assignment ***π***_*t*_, so that the stochastic assignment regularizes the prediction. This joint optimization updates the decoder, phenotype embeddings, selector, static encoder Enc^stat^, and, through **z**_*t*_, the attention module, both encoders, and the input embedding. The anti-collapse objective is applied together with this joint training to prevent the selector from leaving phenotypes unused, affecting the selector and upstream representation but not the phenotype embeddings or the decoder.

##### B.4.3 Anti-collapse objective

The anti-collapse term prevents joint training from leaving some phenotypes unused by encouraging uniform batch-level occupancy, following CAMELOT [6]. Let 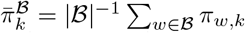 denote the mean assignment mass of phenotype *k* over a mini-batch. The objective is the reverse Kullback–Leibler divergence from the uniform distribution,

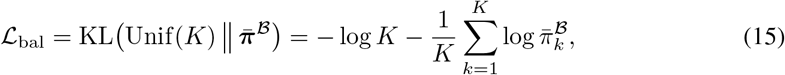

and is applied jointly with the prediction objective. Its gradient with respect to 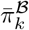 scales as 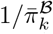, so the restoring force is strongest for a phenotype approaching disuse. The term therefore controls assignment occupancy without changing the geometry of the phenotype embeddings or the decoding function.

#### B.5 Training procedure

Training proceeds in three stages, moving from a *K*-independent longitudinal representation to phenotype assignments and finally to the jointly optimized endpoint prediction model. Stage 1 trains the input embedding, both encoders, and the attention module without phenotype embeddings or other phenotype-specific parameters. Stage 2 uses the resulting contexts to initialize and pretrain the predictive phenotype layer, including the selector and decoder. Stage 3 unfreezes the full model and jointly optimizes the endpoint, forecasting, contrastive, disentanglement, and anti-collapse objectives.

##### Stage 1

Stage 1 trains everything required to produce the context representation **z**_*t*_, with the selector, phenotype embeddings, and decoder excluded because no predictive phenotype layer exists yet. The objective is

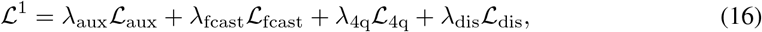

optimized for *E*_1_ epochs, where the loss weights *λ*_aux_, *λ*_fcast_, *λ*_4q_, and *λ*_dis_ are selected on the validation split (Section C.3). The resulting representation is subsequently frozen for phenotype-count selection and provides the fixed context space used to initialize the phenotype embeddings.

##### Stage 2

Stage 2 initializes the predictive phenotype layer from the Stage-1 contexts, one context **z**_*w*_ per window read at its decision point. The phenotype embeddings are initialized as

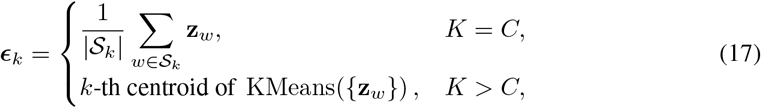

where *S*_*k*_ is the set of training windows whose endpoint label is class *k*, so that at *K* = *C* each phenotype embedding starts at the mean context of its diagnostic class. The selector is then pretrained for *E*_2_ epochs using *L*_sel_ (Section B.4.1) while the phenotype embeddings remain fixed and the Stage-1 representation, attention module, and decoder remain frozen. The pseudo-label 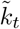 is the endpoint class for *K* = *C* and the nearest phenotype embedding for *K* > *C*, giving the selector an initial mapping from contexts to phenotype assignments before endpoint supervision is introduced. The decoder is then pretrained for *E*_dec_ epochs with *L*_cls_ (Section B.4.2) while the remaining components stay frozen and the model is kept in evaluation mode.

##### Stage 3

Stage 3 unfreezes the full model and jointly optimizes

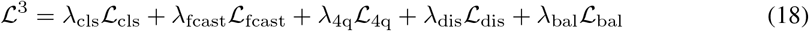

for *E*_3_ epochs, replacing the auxiliary diagnostic objective of Stage 1 by the endpoint objective *L*_cls_. Its weights are again selected on validation, with *λ*_cls_ = 1 fixed to remove a redundant global scale.

##### Optimization and model selection

All stages use the AdamW optimizer. Validation is evaluated every *E*_val_ epochs using the same sampler and batch size as training, and the restored checkpoint is the one maximizing the harmonic mean of validation Purity and macro-F1, both computed at window decision points from the same hard phenotype-to-dominant-class mapping. Training stops after *P* = 10 consecutive validation evaluations without improvement once *E*_min_ = 50 epochs have elapsed.

### C Experiments

#### C.1 Dataset

Experiments use the harmonized ADNIMERGE table, which combines demographic, neuropsychological, genetic, and structural MRI measurements across ADNI-1/GO/2/3. The raw cohort contains 16, 421 visits from 2, 430 participants. The study retains 21 features, comprising 16 longitudinal variables and 5 static covariates. The prediction target is the clinical diagnosis at a future horizon *h*, where *h* ∈ *{*0, 12, 24}, corresponding to current-visit, 1-year-ahead, and 2-years-ahead scenarios, with three classes: cognitively normal (CN), mild cognitive impairment (MCI), and Alzheimer’s Disease (AD). After cohort filtering for genotype availability, visit quality, and a minimum of three visits per participant, the final cohort contains 1,788 participants and 10,941 visits. Table A4 summarizes the retained covariates and their distributions.

Preprocessing followed the process in [5, 13]. Missing longitudinal measurements are handled with participant-level forward filling followed, when necessary, by population-level median or mode imputation, while the missingness mask **m**_*i*_ records which values were originally measured. Patient trajectories are sorted by time and standardized using statistics from the training split only. All splits are performed at the patient level, with 70% of participants used for training, 10% for validation, and 20% for testing, ensuring that no patient’s visits occur across splits.

TRACE constructs longitudinal rolling windows with a window size of *W* = 7. Windows are retained only when at least three real visits are present within the window, excluding padded positions. Endpoint labels are then assigned at the window’s decision point using the specified prediction horizon, so each window represents the history available up to that point while the target corresponds to the subsequent endpoint outcome. The same patient-level split is maintained after window construction, and all preprocessing statistics are fitted on the training patients only.

#### C.2 Evaluation Metrics

All metrics are computed at the window level over the *N*_*W*_ evaluated windows. Each window *w* carries an endpoint label *y*_*w*_∈{1, …, *C}*, the outcome at the prediction horizon. The corresponding hard phenotype assignment is 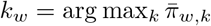, which is used to evaluate the learned phenotype partition. The predicted scores 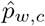 are used for endpoint classification 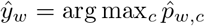. Both evaluation components therefore use the same endpoint label but assess different aspects of the model.

##### C.2.1 Clustering Quality

Clustering quality evaluates how the learned phenotype partition aligns with endpoint labels. Let *P*_*k*_ = {*w* : *k*_*w*_ = *k*} denote the windows assigned to phenotype *k* and *Y*_*c*_ = {*w* : *y*_*w*_ = *c*} the windows with endpoint class *c*.

##### Purity

Purity assigns each phenotype its most frequent endpoint class and reports the fraction of windows that agree with their phenotype’s class,

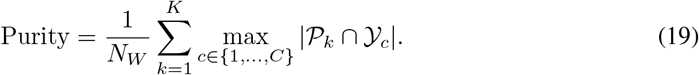

##### Normalized Mutual Information

NMI [16] measures the information the phenotype assignment carries about the endpoint label, normalized by the entropy of the phenotype and diagnostic label assignments,

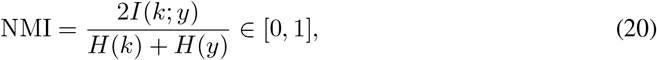

**Table A4:** Summary of the ADNI cohort used in this work (1,788 participants, 10,941 visits). Static covariates are summarized over participants and time-varying covariates over visits. Continuous variables report the mean and range, while categorical variables report the proportion and value of the modal category. Values are shown as recorded, before imputation.

| Group | Variable | Type | Summary |
| --- | --- | --- | --- |
| **Static covariates** |  |  |  |
| Demographic | Race | Cat. | 0.93 (White) |
|  | Ethnicity | Cat. | 0.96 (Not Hisp./Latino) |
|  | Education | Cont. | 16.02 (6–20) |
|  | Marital status | Cat. | 0.76 (Married) |
| Genetic | APOE<sub>4</sub> | Cont. | 0.56 (0–2) |
| **Time-varying covariates** |  |  |  |
| Demographic | Age | Cont. | 73.27 (55–91.40) |
| Biomarker | Entorhinal | Cont. | $3.5 \times 10^3$ ($9.8 \times 10^2 - 7.1 \times 10^3$) |
| | Fusiform | Cont. | $1.7 \times 10^4$ ($7.7 \times 10^3 - 3.0 \times 10^4$) |
| | Hippocampus | Cont. | $6.8 \times 10^3$ ($2.2 \times 10^3 - 1.1 \times 10^4$) |
| | Mid temporal | Cont. | $1.9 \times 10^4$ ($8.0 \times 10^3 - 3.2 \times 10^4$) |
| | Ventricles | Cont. | $4.2 \times 10^4$ ($5.6 \times 10^3 - 1.6 \times 10^5$) |
| | Whole brain | Cont. | $1.0 \times 10^6$ ($5.2 \times 10^5 - 1.5 \times 10^6$) |
| | Intracranial | Cont. | $1.5 \times 10^6$ ($7.2 \times 10^5 - 3.5 \times 10^6$) |
| Cognitive | ADAS-11 | Cont. | 10.77 (0–69) |
|  | ADAS-13 | Cont. | 16.63 (0–84) |
|  | CDR Sum of Boxes | Cont. | 1.92 (0–17) |
|  | Mini-Mental State | Cont. | 26.86 (0–30) |
|  | RAVLT Forgetting | Cont. | 4.16 (0–15) |
|  | RAVLT Immediate | Cont. | 35.61 (0–75) |
|  | RAVLT Learning | Cont. | 4.19 (–5–14) |
|  | RAVLT Percent | Cont. | 56.97 (0–100) |

where *I*(·;·) and *H*(·) are the empirical mutual information and entropy of the phenotype assignments *k*_*w*_ and the endpoint labels *y*_*w*_. It is 0 when phenotypes and labels are independent and 1 when they agree up to relabeling.

##### Adjusted Rand Index

ARI [17] counts window *pairs* placed together or apart by both the partition and the labels, and subtracts the count expected under a random partition with the same phenotype and class sizes. A partition unrelated to the labels therefore scores 0 in expectation at every *K*, and only an exact correspondence between phenotypes and classes scores 1.

##### Silhouette Index

Silhouette Index (SI) is computed using the endpoint-label vectors rather than the feature representation. For each window *w* assigned to phenotype *k*_*w*_, let *a*_*w*_ denote its mean within-phenotype distance and *b*_*w*_ its mean distance to the nearest competing phenotype, where distance is the *L*_1_ distance between the corresponding endpoint label vectors. The silhouette score is

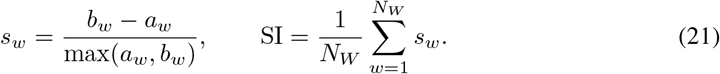

##### C.2.2 Endpoint Classification Performance

Classification performance evaluates the endpoint label *y*_*w*_ from the phenotype-mediated prediction path. Accuracy and macro-F1 use the single operating point *ŷ*_*w*_, whereas macro-AUROC and macro-AUPRC use the class scores 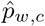 across all thresholds and therefore evaluate ranking performance rather than one fixed decision.

##### Accuracy

Accuracy measures the fraction of windows whose predicted endpoint class matches the true endpoint class,

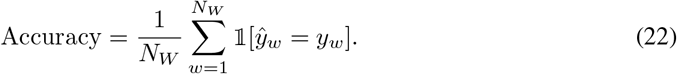

##### Macro-F1

With P_*c*_ and R_*c*_ the one-vs-rest precision and recall of class *c* under the arg max decision, macro-F1 averages the class-wise harmonic mean of precision and recall,

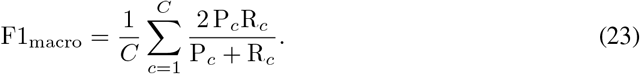

##### Macro-AUROC and macro-AUPRC

For each class *c*, one-vs-rest AUROC is computed from the scores 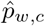 with ties counted as half, and average precision is computed as the step-wise sum of precision increments over the precision–recall curve obtained by thresholding 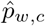 Both are averaged over the *C* classes.

##### Purity and accuracy at *K* = *C*

Purity in Eq. (19) and Accuracy in Eq. (22) are computed on the same windows and the same endpoint labels, and at *K* = *C* they reduce to a single quantity for any method whose endpoint decision is constant within a phenotype. Let *ĉ*(*k*) denote the class such a method predicts for every window assigned to phenotype *k*, so that *ŷ*_*w*_ = *ĉ*(*k*_*w*_). Then

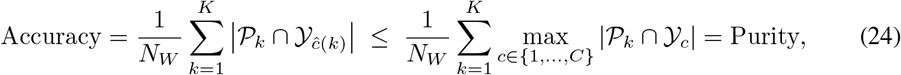

with equality exactly when each phenotype predicts its own most frequent endpoint class. Purity is therefore the accuracy ceiling of a phenotype-constant decision rule rather than an independent measurement, and the ceiling is attained whenever the decoder disagrees with the majority label of no phenotype. This is the situation for the methods of Tables 1 and 2 that predict through the phenotype assignment, namely AC-TPC, KM-E2P(**z**), KM-E2P(*y*), CAMELOT, T-Phenotype at *h* ∈ {0, 12}, and TRACE, whose Purity and Accuracy agree to the reported precision because the two columns evaluate one decision rule from two directions rather than two independent properties of the model. KM-DTW has no predictive head and is scored under the rule stated in the footnote of Table 2. T-Phenotype at *h* = 24 likewise scores above the ceiling of Eq. (24), which is possible only because its predictor is not exactly phenotype-constant at that horizon. Purity is retained for comparability with prior work, but at *K* = *C* it should not be read as evidence separate from accuracy for the phenotype-mediated methods. NMI, ARI, and SI carry the partition-level evidence that accuracy does not contain.

##### Metric behavior at *K* > *C*

A single endpoint class may be intentionally divided across multiple phenotypes to capture within-class progression structure. Purity can therefore remain high or increase under refinement, while NMI and ARI measure continued agreement with the original *C*-class labeling rather than the quality of the additional within-class structure. SI is even more restrictive: splitting a class across phenotypes introduces a competing phenotype with the same endpoint label and can drive its score toward zero even when the resulting phenotypes are internally coherent.

#### C.3 Hyperparameter Search

Hyperparameters are selected independently for each reporting seed using Optuna’s multivariate, grouped Tree-structured Parzen Estimator (TPE). Each trial is trained on the training split and selected using validation macro-AUPRC. Continuous weights and learning rates are sampled in log space, while discrete and categorical parameters use the grids shown in Table A5. For TRACE, *λ*_cls_ = 1 is fixed to remove the redundant global scaling degree of freedom, given a fixed *K* and batch size |*B*|= 64. The embedding type and encoder backbone are evaluated as a separate grid on the most challenging task, and fixed across prediction horizons. The search budget is 50 trials per study, with reduced training budgets during search followed by full-budget retraining of the selected configuration.

#### C.4 Criteria for Choosing *K*

The number of phenotypes *K* is selected on the validation split before Stage 2 and is not jointly optimized with the remaining hyperparameters. Since Stage 1 is independent of *K*, candidate values can be evaluated by clustering the frozen Stage-1 contexts without retraining the representation. Selection is performed separately for each prediction horizon.

The candidate set is {*C* + 1, …, 3*C* + 1}. For each candidate *K, K*-means++ is applied with ten restarts to the validation contexts **z**_*w*_ read at the decision point, giving a candidate assignment *k*_*w*_. Candidate quality is evaluated along two complementary axes: geometric coherence, measured by an *m*-nearest-neighbor silhouette, and alignment with the horizon-specific endpoint labels, measured by adjusted mutual information (AMI).

##### Geometric coherence

Let ^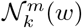^ denote the *m* nearest neighbors of window *w* among the windows assigned to phenotype *k*, and let

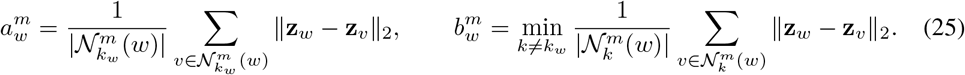

The *m*-nearest-neighbor silhouette *S*^*m*^ is the mean of 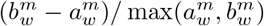 over the validation windows, and the neighborhood size *m* is swept over the considered range as a robustness check.

##### Outcome alignment

The adjusted mutual information between the candidate assignments *k*_*w*_ and the horizon-*h* endpoint labels *y*_*w*_ is

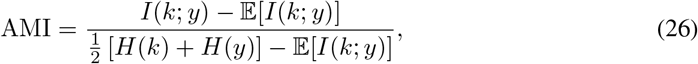

with the expectation taken under random assignments with the same phenotype and class sizes. AMI is preferred to the label-based silhouette of Eq. (21) because the latter favors *K* = *C* when a diagnostic class is split across multiple phenotypes, while unadjusted NMI can increase with *K*.

##### Selection rule

Both scores lie in [−1, 1] and are rescaled to [0, 1] before being combined by their harmonic mean *J*(*K*), so that a candidate must simultaneously exhibit coherent geometry and outcome-relevant structure. The selected phenotype count *K*^*⋆*^ is the smallest maximizer of *J*(*K*) averaged over the reporting seeds. Candidates producing empty phenotypes or phenotypes with fewer than two windows are excluded, and the procedure is repeated independently for each prediction horizon.

#### C.5 Compute Resources

All experiments were run using one NVIDIA Quadro RTX 6000 (24 GB) 8 CPU cores, and 32 GB of host memory per job. Given a fixed hyperparameter configuration, each TRACE run required approximately 40 minutes per seed, while AC-TPC and CAMELOT required approximately 30–35 minutes, KM-E2P approximately 20 minutes, KM-DTW approximately 10 minutes, and T-Phenotype approximately 3 hours. All reported results use ten random seeds. Hyperparameters were selected independently per seed using 50 Optuna TPE trials with reduced training budgets, followed by full-budget retraining of the selected configuration. The *K*-selection sweep reused cached Stage-1a representations rather than retraining the model for each candidate *K*.

### D Additional Results

#### D.1 Phenotype Discovery *K* > *C*

The *K* > *C* analysis of Section 5 uses *K* = 4 at *h* = 24 months, where the discovered partition contains 178, 202, 158, and 87 windows for *k*_0_–*k*_3_, compared with 221, 257, and 147 windows for the label-matched partition. Figure A1 provides a complementary view of the discovered phenotypes by showing how their mean feature values evolve over the history window, with the open circle marking the beginning and the arrowhead the decision point. The trajectories reveal distinct temporal patterns across the phenotypes: *k*_0_ remains relatively compact across most feature pairs, whereas *k*_1_ occupies a different region and undergoes larger changes, particularly across cognitive and memory-related variables. The newly discovered *k*_3_ generally lies between *k*_0_ and *k*_1_ in the joint feature space, sharing characteristics with both phenotypes, but follows its own trajectory rather than reproducing the evolution of either one. In several cognitive–structural planes, *k*_3_ remains intermediate between the two phenotypes while showing a distinct direction or magnitude of change. For example, its hippocampal, fusiform, and entorhinal trajectories remain closer to *k*_0_, whereas its cognitive trajectories tend to occupy an intermediate position between *k*_0_ and *k*_1_. Its evolution is also distinct in the RAVLT and ICV planes, where the trajectory differs more clearly from those of both *k*_0_ and *k*_1_. Thus, *k*_3_ is not simply a subdivision or interpolation of the existing phenotypes, but an intermediate phenotype characterized by a distinct combination and temporal evolution of cognitive, memory, and structural features. These qualitative patterns are consistent with, rather than independent confirmation of, the metric behavior at *K* > *C* discussed in Section C.2.

#### D.2 Broader Impacts

TRACE is designed as a general framework for longitudinal multiclass classification settings in which heterogeneity in how observations evolve over time may be important for timely decision making. When the possible outcomes are known but the ways in which individuals can progress toward those outcomes are less well understood, TRACE can provide an overview of the distinct temporal phenotypes present in the data and characterize the visits and variables most associated with each phenotype through its saliency maps. Such a representation may help domain experts identify different progression pathways and determine which features or time points warrant closer attention as an individual’s trajectory evolves. At the same time, the discovered phenotypes and their saliency patterns should be viewed as data-driven hypotheses rather than established mechanisms or clinical subtypes. Expert assessment is needed to determine whether the identified pathways and salient features are meaningful in the relevant application domain, and external validation is needed to establish their reproducibility and generalizability across populations and settings.

**Figure A1:**
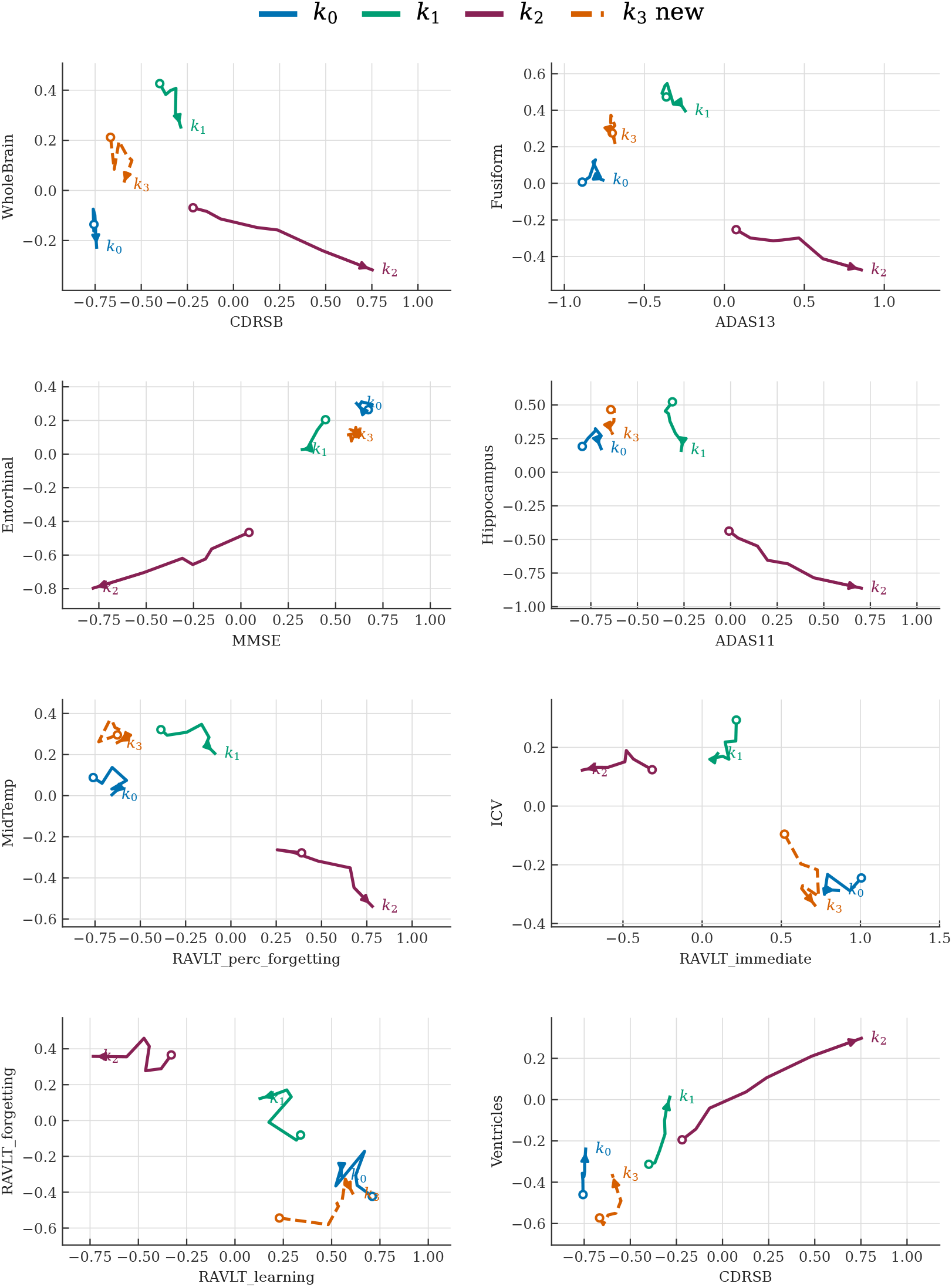
Mean feature-space trajectories of the four phenotypes discovered at *K* = 4 *> C* on ADNI at *h* = 24 months. Each panel plots the phenotype-mean trajectory through the plane spanned by two standardized clinical variables, averaged over the windows assigned to that phenotype (178, 202, 158, and 87 windows for *k*_0_–*k*_3_). The open circle marks the beginning of the history window and the arrowhead the decision point. The feature trajectories show that the newly discovered *k*_3_ generally lies between *k*_0_ and *k*_1_, while following a distinct temporal path. Across several planes, *k*_3_ shares structural characteristics with *k*_0_ but exhibits cognitive and memory profiles intermediate between *k*_0_ and *k*_1_ with distinct evolution paths.

**Table A5:**
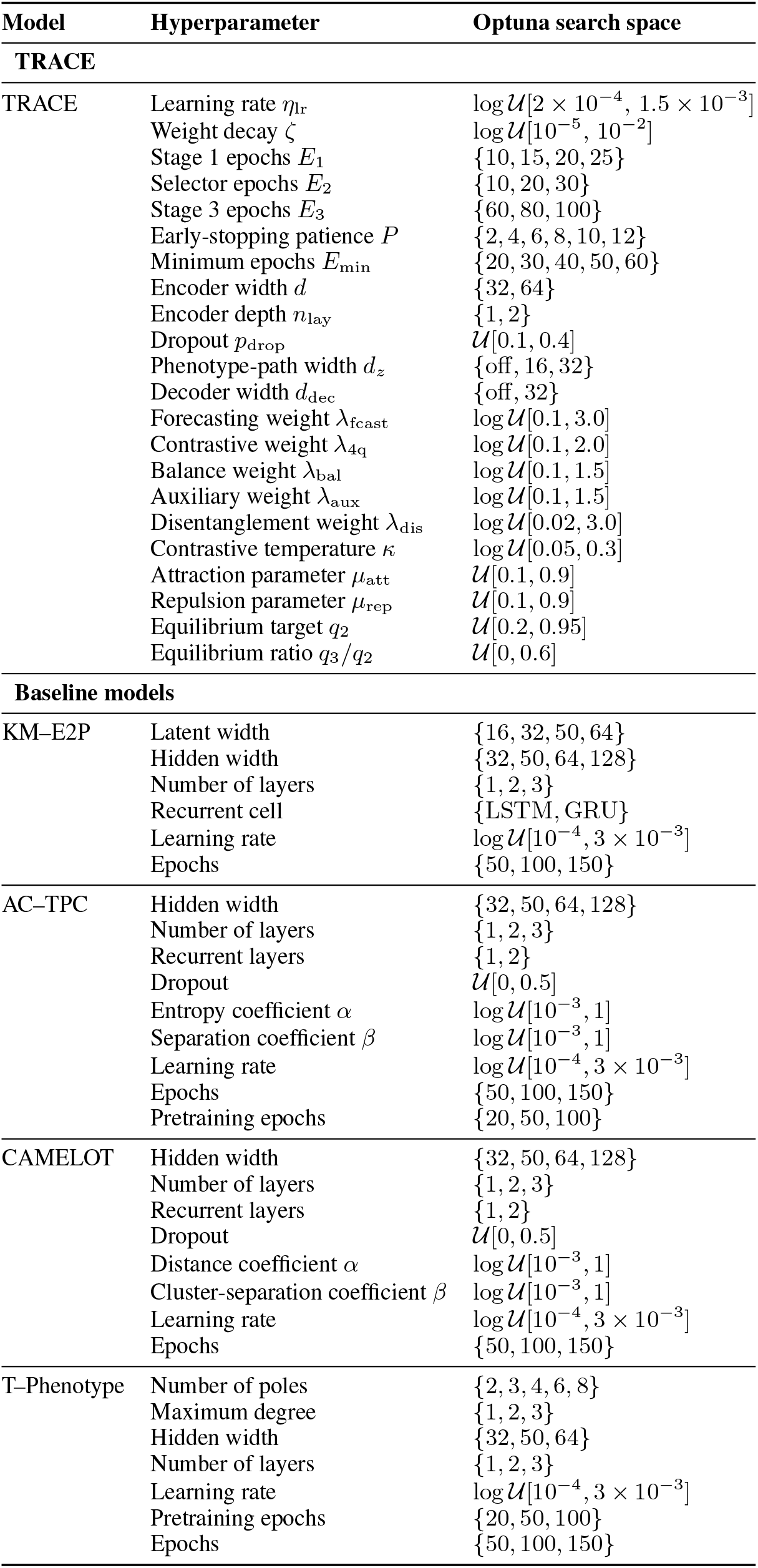
Hyperparameter search spaces used with Optuna. Each row lists the parameters searched for a given model and the corresponding search space. The TRACE configuration searches the full space. Baselines use their own method-specific search spaces.

## References

[1] Menglin Lu, Yujie Zhang, Suixia Zhang, Hanrui Shi, and Zhengxing Huang. Knowledge-aware patient representation learning for multiple disease subtypes. Journal of Biomedical Informatics, 138:104292, 2023.

[2] Benjamin Shickel, Patrick James Tighe, Azra Bihorac, and Parisa Rashidi. Deep ehr: a survey of recent advances in deep learning techniques for electronic health record (ehr) analysis. IEEE journal of biomedical and health informatics, 22(5):1589–1604, 2017.

[3] Zhengping Che, Sanjay Purushotham, Kyunghyun Cho, David Sontag, and Yan Liu. Recurrent neural networks for multivariate time series with missing values. Scientific reports, 8(1):6085, 2018.

[4] Edward Choi, Mohammad Taha Bahadori, Jimeng Sun, Joshua Kulas, Andy Schuetz, and Walter Stewart. Retain: An interpretable predictive model for healthcare using reverse time attention mechanism. Advances in neural information processing systems, 29, 2016.

[5] Changhee Lee and Mihaela Van Der Schaar. Temporal phenotyping using deep predictive clustering of disease progression. In International conference on machine learning, pages 5767–5777. PMLR, 2020.

[6] Henrique Aguiar, Mauro Santos, Peter Watkinson, and Tingting Zhu. Learning of cluster-based feature importance for electronic health record time-series. In International conference on machine learning, pages 161–179. PMLR, 2022.

[7] Mohammad Al Olaimat, Serdar Bozdag, and Alzheimer’s Disease Neuroimaging Initiative. Ta-rnn: an attention-based time-aware recurrent neural network architecture for electronic health records. Bioinformatics, 40(Supplement_1):i169–i179, 2024.

[8] Yuchao Qin, Mihaela van der Schaar, and Changhee Lee. T-phenotype: Discovering phenotypes of predictive temporal patterns in disease progression. In Proceedings of The 26th International Conference on Artificial Intelligence and Statistics, volume 206 of Proceedings of Machine Learning Research, pages 4605–4624. PMLR, 2023.

[9] Weijieying Ren, Jingxi Zhu, Zehao Liu, Tianxiang Zhao, and Vasant Honavar. A comprehensive survey of electronic health record modeling: From deep learning approaches to large language models. arXiv preprint arXiv:2507.12774, 2025.

[10] Jacob W Vogel, Alexandra L Young, Neil P Oxtoby, Ruben Smith, Rik Ossenkoppele, Olof T Strandberg, Renaud La Joie, Leon M Aksman, Michel J Grothe, Yasser Iturria-Medina, et al. Four distinct trajectories of tau deposition identified in alzheimer’s disease. Nature medicine, 27(5):871–881, 2021.

[11] Su Xian, Monika E Grabowska, Iftikhar J Kullo, Yuan Luo, Jordan W Smoller, Theresa L Walunas, Wei-Qi Wei, Gail P Jarvik, Sean D Mooney, and David R Crosslin. Transformer patient embedding using electronic health records enables patient stratification and progression analysis. npj Digital Medicine, 8(1):521, 2025.

[12] Ronald Carl Petersen, Paul S Aisen, Laurel A Beckett, Michael C Donohue, Anthony Collins Gamst, Danielle J Harvey, Clifford R Jack Jr, William J Jagust, Leslie M Shaw, Arthur W Toga, et al. Alzheimer’s disease neuroimaging initiative (adni) clinical characterization. Neurology, 74(3):201–209, 2010.

[13] Shahriar Noroozizadeh, Jeremy C Weiss, and George H Chen. Temporal supervised contrastive learning for modeling patient risk progression. In Machine Learning for Health (ML4H), pages 403–427. PMLR, 2023.

[14] Hiroaki Sakoe and Seibi Chiba. Dynamic programming algorithm optimization for spoken word recognition. IEEE transactions on acoustics, speech, and signal processing, 26(1):43–49, 1978.

[15] Romain Tavenard, Johann Faouzi, Gilles Vandewiele, Felix Divo, Guillaume Androz, Chester Holtz, Marie Payne, Roman Yurchak, Marc Rußwurm, Kushal Kolar, et al. Tslearn, a machine learning toolkit for time series data. Journal of machine learning research, 21(118):1–6, 2020.

[16] Nguyen Xuan Vinh, Julien Epps, and James Bailey. Information theoretic measures for clusterings comparison: is a correction for chance necessary? In Proceedings of the 26th annual international conference on machine learning, pages 1073–1080, 2009.

[17] Lawrence Hubert and Phipps Arabie. Comparing partitions. Journal of classification, 2(1): 193–218, 1985.

[18] Peter J Rousseeuw. Silhouettes: a graphical aid to the interpretation and validation of cluster analysis. Journal of computational and applied mathematics, 20:53–65, 1987.

[19] Yury Gorishniy, Ivan Rubachev, and Artem Babenko. On embeddings for numerical features in tabular deep learning. Advances in Neural Information Processing Systems, 35:24991–25004, 2022.

[20] Seyed Mehran Kazemi, Rishab Goel, Sepehr Eghbali, Janahan Ramanan, Jaspreet Sahota, Sanjay Thakur, Stella Wu, Cathal Smyth, Pascal Poupart, and Marcus Brubaker. Time2vec: Learning a vector representation of time. arXiv preprint arXiv:1907.05321, 2019.

[21] Pengyu Cheng, Weituo Hao, Shuyang Dai, Jiachang Liu, Zhe Gan, and Lawrence Carin. Club: A contrastive log-ratio upper bound of mutual information. In International conference on machine learning, pages 1779–1788. PMLR, 2020.

[22] Prannay Khosla, Piotr Teterwak, Chen Wang, Aaron Sarna, Yonglong Tian, Phillip Isola, Aaron Maschinot, Ce Liu, and Dilip Krishnan. Supervised contrastive learning. Advances in neural information processing systems, 33:18661–18673, 2020.

[23] Aaron van den Oord, Yazhe Li, and Oriol Vinyals. Representation learning with contrastive predictive coding. arXiv preprint arXiv:1807.03748, 2018.

[24] David M. Blei, Andrew Y. Ng, and Michael I. Jordan. Latent Dirichlet allocation. Journal of Machine Learning Research, 3:993–1022, 2003.

